# Prevalence of dementia among community-dwelling older adults in rural Uganda – findings from DEPEND Uganda (Dementia Epidemiology, unmet need and co-developing solutions in Uganda)

**DOI:** 10.64898/2026.09.18.26363378

**Authors:** Josephine E Prynn, Racheal Alinaitwe, Beatrice Kimono, Ronald Makanga, Michael Mubiru, Noeline Nakasujja, Tunde Peto, Claire Steves, Joseph Mugisha, Martin Prince

**Affiliations:** King’s College London; MRC, UVRI & LSHTM Uganda Research Unit; Makerere University; Queen’s University Belfast

## Abstract

**INTRODUCTION:** The number of people living with dementia in many African countries is anticipated to treble by 2050. However, data from Uganda are scarce, with no published prevalence studies using a comprehensive approach to dementia diagnosis.

We used methods developed by the 10/66 Dementia Research Group to estimate dementia prevalence in rural South-West Uganda, and its association with socio-demographics and functional status.

**METHODS:** The cross-sectional study was nested within the General Population Cohort. In Phase 1, all cohort participants aged 60+ underwent cognitive screening. In Phase 2, we invited all those with impaired cognition, 50% with mildly impaired cognition, and 15% with unimpaired cognition for assessment. We also assessed functional status using WHODAS-12, IDEA-IADL and reported care needs.

**RESULTS:** Over 80% of eligible participants underwent screening, and of those sampled, over 90% were assessed. Dementia prevalence was 18.6%, and most cases were mild or very mild. Dementia was associated with increased age and lower educational attainment. People with dementia had lower performance at IADLs, but similar levels of disability and care needs.

**DISCUSSION:** Dementia prevalence in Uganda is high, affecting nearly 1 in 5 people aged 60 and over. This is higher than most other estimates from the region, particularly others that have used two-phase approaches and clinical DSM diagnoses, which may under-estimate dementia prevalence due to attrition between study phases and cultural variations in reporting functional impairment by informants. Any planned health and social care services for older people should include support for people with dementia and their families.

## Introduction

The number of people living with dementia is increasing globally, with the most rapid changes predicted in countries in the lowest income brackets. Strikingly, in many African countries the number is expected to triple between 2019 and 2050, due to a combination of population ageing and growth.[1]

However, these estimates are based on limited data, as relatively few studies examining dementia prevalence have been conducted in African settings, and many have been small and without in-depth cognitive assessment.[2] While there is a relative wealth of prevalence data from other global regions, extrapolation of these findings to different contexts should be done with caution, as exposures that accumulate over a lifetime will vary substantially between settings. Furthermore, there are diagnostic considerations to be made in many lower income settings, such as tailoring cognitive assessments to lower literacy populations and specific cultural norms.

Understanding the current burden of dementia, who is most at risk, and how dementia affects a person’s ability to function is crucial for health and social care systems to be able to plan how best to support people with dementia and their families both now and in the future. A systematic review of dementia prevalence in Africa in 2017 found that there had been 12 studies published, of which one was in East Africa, and none were in Uganda.[3] Overall prevalence in Africa was estimated at 1.74% in adults aged 60-64 rising to 23.31% in adults aged 85 and older. There have since been three studies published on prevalence of cognitive impairment or dementia in Ugandan populations.[4–6] All were population-based studies in rural areas but used different approaches to estimate dementia prevalence. Using the Community Screening Instrument for Dementia (CSI-D) in South-West Uganda, Mubangizi et al estimated prevalence of probable dementia at 20.0%.[5] In Northern Uganda, Benyumiza et al used the Brief Cognitive Score for Dementia with back-weighting to assumed sensitivity and specificity values, and estimated prevalence of probable or possible dementia at 23.0%.[4] Wandera et al’s study in Eastern Uganda used the Intervention for Dementia in Elderly Africans (IDEA) cognitive screening tool combined with the IDEA-IADL tool to assess function, and estimated that 20.6% had neurocognitive impairment and 12.0% had dementia.[6] While these studies are helpful in providing an indicator of the burden of cognitive impairment and dementia, all the estimates were based on brief screening or cognitive assessment tools, rather than a comprehensive diagnostic approach to ascertaining dementia status.

Best practice clinical assessment for dementia diagnosis includes cognitive testing, clinical examination and informant interviews.[7] Reproducing this approach in a population study setting to estimate prevalence is a challenge, particularly in low-income and low-literacy settings.

Dementia assessments must be reasonably brief and reproducible by non-experts, and not overly reliant on cognitive assessments that are highly influenced by educational attainment or literacy.[8] In response to this challenge, the 10/66 Dementia Research Group developed an approach to dementia diagnosis using recognised standardised tools (cognitive tests, an informant interview, and a clinical mental state examination) combined algorithmically to generate an output of probable dementia, which was validated against a clinician DSM-IV diagnosis in multiple low- and middle-income settings, and demonstrated a 94% sensitivity and 85-97% specificity for dementia.[8]

This paper uses the approach developed by the 10/66 Dementia Research Group to describe the prevalence of probable dementia in a rural population in Uganda, and its associations with socio-demographic characteristics and disability.

## Methods

### Setting and population

Uganda has a population of nearly 50 million people with over 65 recognised tribes and languages, the largest of which is Baganda, comprising around 15% of the population.[9] The national life expectancy at birth is 68.2 years,[9] and at age 60 is 19.5 years,[10] and approximately 5% of the population is aged 60 or over.[9] An estimated 74% of the population live in rural areas, and 33% of households are engaged in subsistence farming.[9] Institutional care in older age is rare, so the overwhelming majority of older people are community-based.

This study is nested in the General Population Cohort (GPC), a population cohort run by the MRC/UVRI & LSHTM Uganda Research Unit, comprising everyone within a defined area within Kyamulibwa sub-county and Town Council of Kalungu District in South-West Uganda (**Figure 1**), including over 1,500 adults aged 60+.[11] Kyamulibwa is a rural subsistence farming area, and most residents have Luganda as their first language.

**Figure 1.**
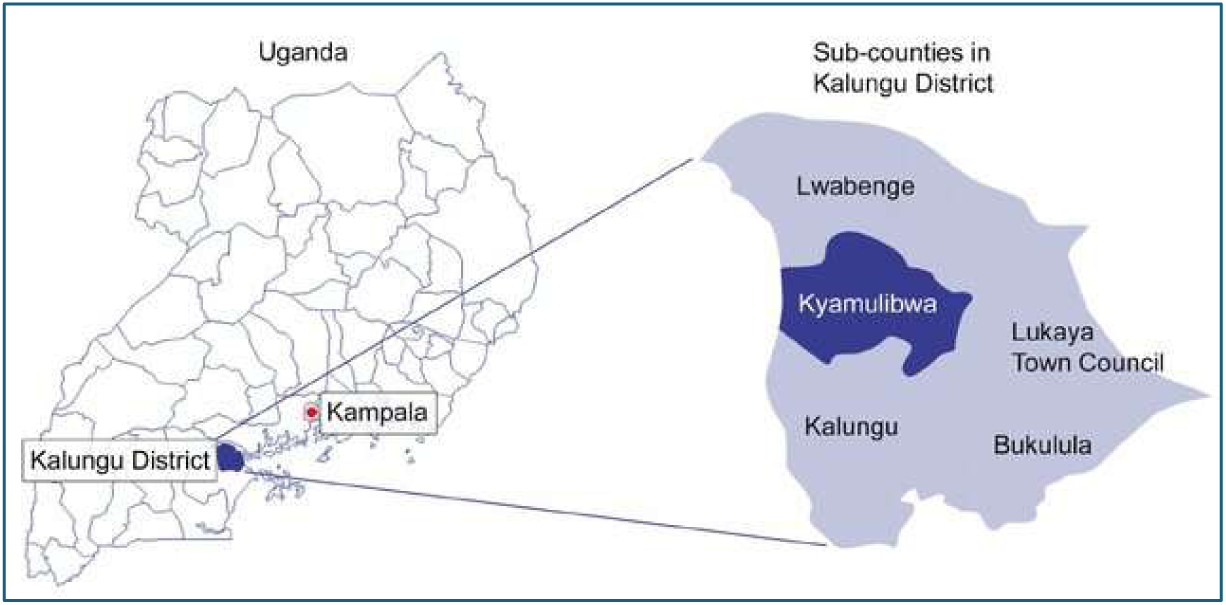
Map of study area

The GPC was set up in 1989 originally as an HIV surveillance cohort, and has since expanded to include research on other communicable diseases, non-communicable disease, mental health and disability. It includes 25 core villages and one pilot village. Every one to two years since its inception the GPC population has been enumerated through a comprehensive census. Following the census, all adults are invited to participate in the GPC medical survey, where data are collected using a structured questionnaire, anthropometry, and a point-of-care HIV test. Further details of the GPC processes are described elsewhere.[11]

### Study design overview

This was a two-phase cross-sectional prevalence study, outlined in **Figure 2**. Phase 1 was integrated into the Round 28 of the GPC medical survey, when all GPC participants aged 60+ underwent brief cognitive screening.

**Figure 2.**
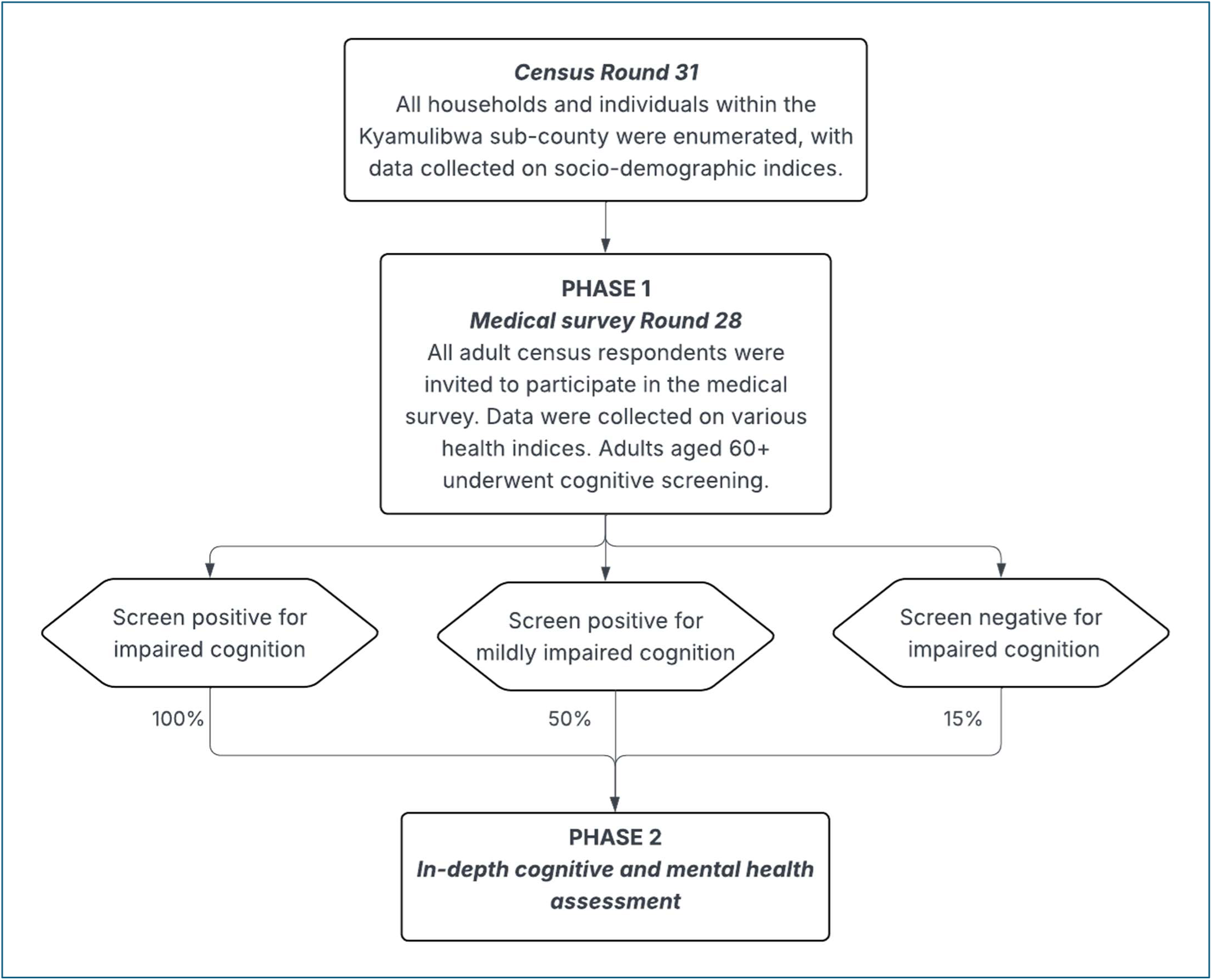
Study processes

For Phase 2, participants were sampled based on their screening results: all those with impaired cognition, 50% of those with mildly impaired cognition, and 15% of those with unimpaired cognition were invited for in-depth cognitive and mental health assessment, as described in detail below.

Each participant was also asked to identify an informant to be interviewed - their main caregiver if they had one, or someone who knew them well.

Finally, all sampled participants and informants were invited to attend the GPC clinic to undergo further questionnaires and investigations.

### Phase 1: Screening

#### Cognitive screening tools and cut-points for impaired cognition

The cognitive screening consisted of:

• 10-word recall (immediate and delayed recall of 10 familiar words) [12]

• Animal naming (listing of as many animals as possible within 1 minute) [13]

• Digit span (immediate recall of an increasing list of numbers going forwards and then backwards) [14]

We chose these screening assessments as they had been used in the Wellbeing of Older People’s Study (WOPS) in an overlapping geographical region of Uganda between 2009 and 2020. WOPS was part of the World Health Organisation Study on global AGEing and adult health (SAGE).[15] Analysis of WOPS data allowed us to estimate population normative data for these screening tools and identify population-relevant cut-points for impaired and mildly impaired cognition.

Within the WOPS dataset, we generated a cognitive screening score for each participant, by generating z-scores for each of the three separate cognitive screening components, then calculating the mean of the component scores for each participant. WOPS participants were not representative of the socio-demographic distribution of the underlying population, so we adjusted the mean and standard deviation (SD) of the WOPS cognition scores to the age and education distribution of the GPC census population using inverse probability weighting. For DEPEND, we defined cognitive impairment at screening as a score lower than 2.5 standard deviations below the adjusted WOPS mean, and mild cognitive impairment at screening as between 1 and 2.5 standard deviations below the adjusted WOPS mean.

#### Screening procedures

For 20 of the 26 villages, we sampled participants for Phase 2 immediately following their cognitive screening in the GPC Medical Survey Round 28, which followed Census Round 31. To enable sampling for Phase 2 to fall within the necessary timeframe, older people in the remaining 6 villages were screened separately from the routine GPC Round 28/Census Round 31 data collection processes. The sampling frame for these villages was generated from the Census round 30 (2021 – 2023) and one interviewer visited all identified people at home and performed cognitive screening and questions about key dementia risk factors only. They were later also seen in the GPC Medical Survey Round 28, with the cognitive screening stage omitted.

### Phase 2: In-depth cognitive and mental health assessment

Participants were approached at home by an interviewer fluent in Luganda, and invited to participate in Phase 2. The team underwent training in assessing capacity to give informed consent, and used a University of California, San Diego Brief Assessment of Capacity to Consent (UBACC) form to help assess potential participants’ understanding of the information sheets. For people with capacity to consent, informed consent was sought and documented with a signature, or a thumb print witnessed by someone independent of the study. For people without capacity to consent, we sought assent from a close relative, who completed an informant declaration form.

Participants were only included if they gave no verbal or non-verbal communication of refusal to participate in the study. For each participant, we also sought an informant – somebody who knew them well - to answer questions about the participant’s cognition and function.

We used the cognitive and mental health assessment tools required for the 10/66 diagnostic approach to dementia. The tools used are:

**1) Community screening instrument for dementia (CSI-D) [16**]

- **Cognitive assessment component -** A structured assessment of cognition across multiple domains. It has been validated as one of the cognitive assessment tools least biased by educational background.[8]
- **Informant component** - A structured questionnaire for informants about the participant’s cognition and function.
**2) 10-word recall [17, 18]**

Immediate and delayed recall of 10 familiar words in Luganda.
**3) Geriatric mental state (GMS) [19–21]**

A. detailed semi-structured interview assessing for symptoms of depression, anxiety and psychosis, as well as dementia or delirium.

These tools underwent a rigorous translation process. First the tools were translated from English to Luganda, then independently back-translated into English by a second translator. We then addressed any discrepancies between the original and back-translated English versions in a consensus meeting. Finally, the translated versions were shared and discussed within the team of Luganda- and English-speaking interviewers, and further consensus was sought on the most appropriate translations. This updated version was then piloted with a small number of older people to ensure that the questions were clear and well-understood.

For participants with impaired hearing, we provided noise amplifying headphones (Amplicomms TV2500 Headset) to enhance their ability to participate meaningfully in the study.

Data were entered directly onto Redcap on tablet computers.

### Further data collection procedures

Participants were invited with their informant (or a new informant, if the first informant was unavailable) to the study clinic on a separate day for further data collection procedures. At this stage, participants completed the WHO Disability Assessment Schedule short version (WHODAS-12). Informants were asked to complete an identical questionnaire as a proxy for the participant.

Interviewer judgement on the reliability of the participant’s and informant’s responses was recorded at the time of the interview, and if there was doubt about the reliability of the participant’s responses, or the participant could not answer at all, the proxy answers were taken in their place.

The informant was also questioned by the interviewer on the participant’s independence in activities of daily living and care needs, using a series of prompts to encourage descriptive answers. This was summarised by the interviewer as “needs care much of the time”, “needs care occasionally”, or “does not need care, can do everything independently”. The informant also answered questions on the participant’s ability to perform instrumental activities of daily living (IADLs), using the IDEA-IADL tool.[22]

### Analysis

First, we reviewed the socio-demographic characteristics of the population aged 60+ using data from the Census Round 31 for 20 of the villages, and Round 30 for the remaining 6 villages. To characterise socio-economic status (SES), we used a Mokken analysis to generate a unidimensional, monotonic and hierarchical scale using data about the building materials of participants’ homes, ownership of durable assets and access to certain conveniences (e.g. electricity). Details are in **Supplementary Table 1**.

For participants who participated in Phase 2, we used the 10/66 algorithm to identify participants with probable dementia. The 10/66 methodology is described in detail elsewhere,[8] and the algorithm code is available to download at 1066.alzint.org. In brief, the cognitive element of the CSI-D, combined with the standardised 10-word recall test result, generates an output called COGSCORE, and the informant element generates the output RELSCORE. The GMS output is analysed using the AGECAT algorithm which generates symptom clusters, then a score for each clinical syndrome (depression, anxiety, psychosis, and organicity), and finally identifies the syndrome of most clinical importance in that participant. COGSCORE, RELSCORE and the GMS output are combined using the 10/66 diagnostic algorithm, which also accounts for age and educational attainment, to give an output of “probable dementia” or not. Any participants unable to identify an informant were missing the RELSCORE component. For these participants, we used multiple imputation to generate a range of possible RELSCOREs for each participant, based on their census and screening data available, and used the mean of their imputed scores to feed into the 10/66 algorithm. We calculated prevalence of probable dementia for each cognitive screening group, then combined the results accounting for survey design by using inverse probability weights based on their sampling probability with the *svyset* command in Stata.

We used the following approaches to generate a prevalence estimate that accounted for the whole population, including those with missing data. First, we assessed the characteristics of participants sampled for Phase 2 who did not participate. We then used multiple imputation chained equations (MICE) to impute their likely dementia status, based on their characteristics from the census and Phase 1. Next, as this survey was nested within a fully enumerated population, we were able to assess the characteristics of census participants who did not participate in the screening phase (Phase 1) and identify any groups of participants underrepresented in screening. We then extended our use of inverse-probability weighting to estimate the prevalence of dementia when applied to the whole population, as well as accounting for the survey design. To do this, we adjusted the *svyset* command to incorporate both the survey design and any differential drop-out by key characteristics between the census and Phase 1.

As we could not be certain whether data were missing at random (MAR) or missing not at random (MNAR), we also estimated prevalence using a complete case analysis approach as a sensitivity analysis.

To assess the severity of dementia identified, we calculated the Clinical Dementia Rating (CDR), using items extracted from the CSI-D, GMS, and 10-word recall,[23] and assessed for associations between sociodemographic indicators and probable dementia, using logistic regression accounting for the complex survey design. As sensitivity analyses, we also ran these models using dementia with CDR score of 1+ as an outcome, as a potentially more specific indicator of dementia, and then using the dataset that included imputed outcome measures.

Finally, we assessed disability and functional status by probable dementia status. As WHODAS-12 and IDEA-IADL are count data with a highly skewed distribution, we used negative binomial regression analysis to assess the relationship between probable dementia and disability and instrumental activities of daily living (IADLs), using the WHODAS-12 score and IDEA-IADL score respectively. We used ordinal logistic regression to compare the proportion of people whose informants reported needing care much of the time, occasionally, and not at all, which would suggest a reduction in independence in their activities of daily living (ADLs).

## Results

Of the 1615 people aged 60 and over within the census, 1353 (83.8%) completed cognitive screening at Phase 1. Of these, we sampled 324 individuals based on their cognitive screening status, and of these, 294 (90.7%) were seen in the Phase 2, as shown in **Figure 3**.

**Figure 3.**
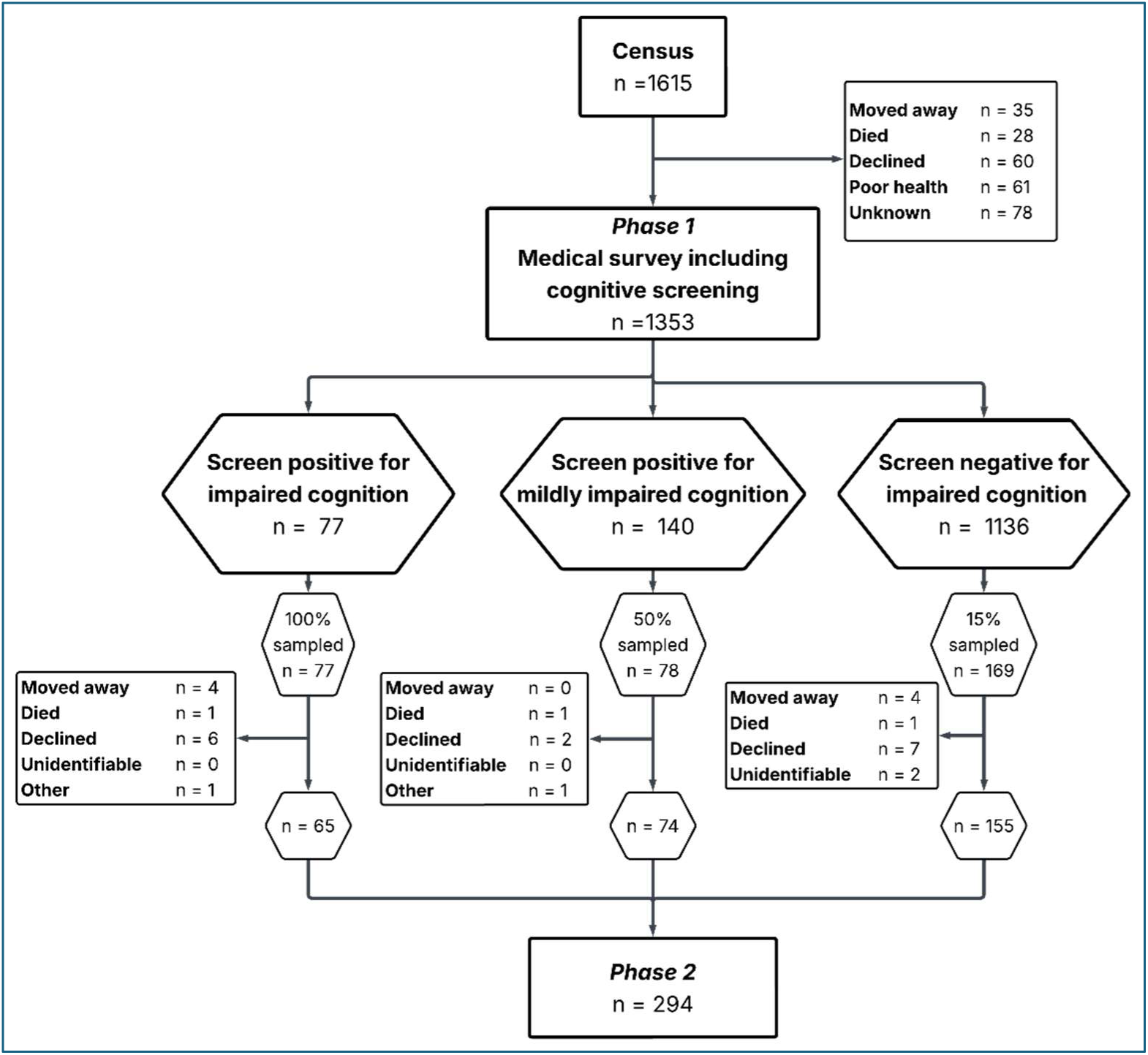
Flow chart of participation

Participation rates at screening were similar among men and women, but the oldest age groups were less likely to participate than the younger old age groups (p<0.001), shown in **Supplementary Table 2**. At Phase 2, participation was more similar across age groups, with slight over-representation of the 70-79 age group compared to the younger or older groups, shown in **Supplementary Table 3**. Across both phases, participation was similar across socio-economic and educational level strata.

As shown in **Table 1**, within the census, 984 (60.9%) of the population were women, and most were in the 60-69 age group. The mean age was 69.7 years overall, 69.4 years in women, and 68.7 years in men. The levels of formal education were low: 20.6% of women and 7.5% of men had received no formal education, and less than 20% of the population had received any secondary education. Most of the participants (84.3%) were subsistence farmers.

**Table 1.** Socio-demographics of the census population.

| **Socio-demographic characteristics** |  | **Women**<br>**n (%)**<br>**n = 984** | **Men**<br>**n (%)**<br>**n = 631** | **Total**<br>**n (%)**<br>**n = 1615** |
| --- | --- | --- | --- | --- |
| **Age group** | 60-69 | 574 (58.3) | 394 (62.4) | 968 (59.9) |
|  | 70-79 | 260 (26.4) | 157 (24.9) | 417 (25.8) |
|  | 80-89 | 124 (12.6) | 68 (10.8) | 192 (11.9) |
|  | 90+ | 26 (2.6) | 12 (1.9) | 38 (2.4) |
| **Formal education** | Nil | 203 (20.6) | 47 (7.5) | 250 (15.5) |
|  | Less than primary | 491 (49.9) | 248 (39.3) | 739 (45.8) |
|  | Completed primary | 147 (14.9) | 163 (25.8) | 310 (19.2) |
|  | Less than secondary | 91 (9.3) | 82 (13.0) | 173 (10.7) |
|  | Completed secondary | 29 (3.0) | 62 (9.8) | 91 (5.6) |
|  | Tertiary | 23 (2.3) | 29 (4.6) | 52 (3.2) |
| **Religion** | Catholic | 608 (61.8) | 361 (57.2) | 969 (60.0) |
|  | Other Christian | 139 (14.1) | 92 (14.6) | 231 (14.3) |
|  | Muslim | 237 (24.1) | 178 (28.2) | 415 (25.7) |
| **Main household income\*** | Subsistence farming | 841 (85.5) | 519 (82.4) | 1360 (84.3) |
|  | Commercial farming | 46 (4.7) | 55 (8.7) | 101 (6.3) |
|  | Wage or salary from job | 23 (2.3) | 19 (3.0) | 42 (2.6) |
|  | Earnings from selling or trading products | 32 (3.3) | 14 (2.2) | 46 (2.9) |
|  | Remittances | 30 (3.1) | 10 (1.6) | 40 (2.5) |
|  | Other | 12 (1.2) | 13 (2.1) | 25 (1.5) |
| **Socioeconomic status\*\*** | 1 (lowest) | 226 (23.0) | 153 (24.3) | 379 (23.5) |
|  | 2 | 154 (15.7) | 86 (13.6) | 240 (14.9) |
|  | 3 | 351 (35.7) | 187 (29.6) | 538 (33.3) |
|  | 4 (highest) | 253 (25.7) | 205 (32.5) | 458 (28.4) |
| * Missing data for one participant<br>**Socio-economic status defined based on responses to a hierarchical scale of questionnaire items, identified using Mokken analysis |  |  |  |  |

We found that after accounting for survey design and missing data, 18.6% (95% CI 14.2% - 22.9%) of the population had probable dementia based on 10/66 criteria, shown in **Table 2**. Our sensitivity analysis using a complete case analysis demonstrated a similar prevalence of 17.4% (95% CI 13.6% - 22.0%), shown in **Supplementary Table 4**.

**Table 2.**
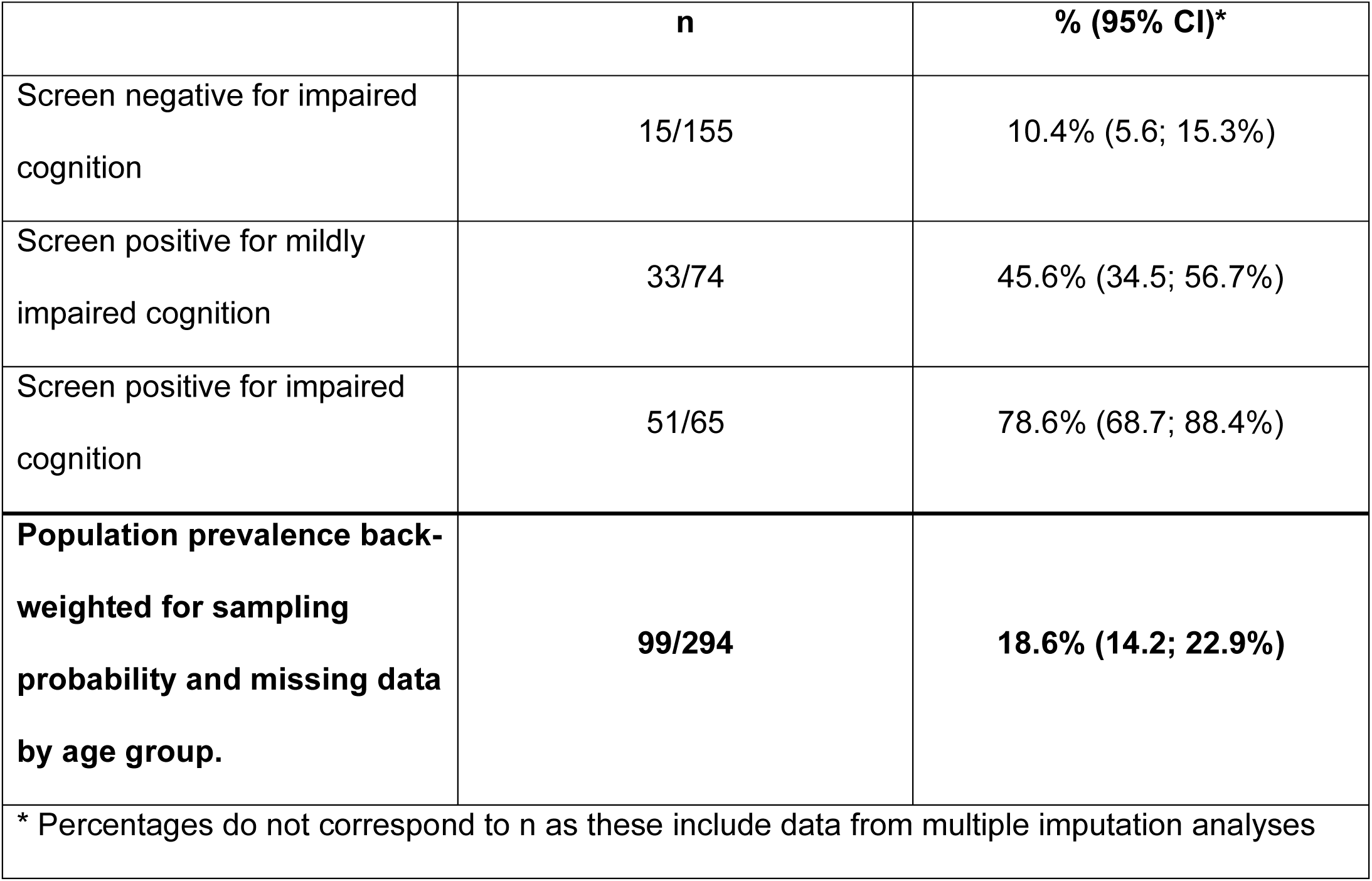
Prevalence of 10/66 dementia within each cognitive screening bracket and overall for adults aged 60 and over.

|  | **n** | **% (95% CI)\*** |
| --- | --- | --- |
| Screen negative for impaired cognition | 15/155 | 10.4% (5.6; 15.3%) |
| Screen positive for mildly impaired cognition | 33/74 | 45.6% (34.5; 56.7%) |
| Screen positive for impaired cognition | 51/65 | 78.6% (68.7; 88.4%) |
| **Population prevalence back-weighted for sampling probability and missing data by age group.** | **99/294** | **18.6% (14.2; 22.9%)** |
| * Percentages do not correspond to n as these include data from multiple imputation analyses |  |  |

The severity of dementia was generally low, with 44.5% of those with probable dementia scoring 0.5 on the Clinical Dementia Rating (CDR) scale, indicating very mild or questionable dementia, and another 43.4% scoring 1, indicating mild dementia. Only 5.1% had a score of 2 (moderate dementia) and we saw no cases with CDR 3 (severe dementia). As a sensitivity analysis, we also calculated prevalence of dementia including only those with CDR of 1 or more, which was 8.8% (95% CI 5.9% - 11.7%).

Prevalence of dementia increased substantially with increasing age, with 7.6% of 60-69 year olds, 19.4% of 70-79 year olds, and 44.4% of people 80 and over affected (p<0.001; d.f. 291), shown in **Table 3**. Formal education was inversely associated with dementia, with an odds ratio (OR) of 0.36 for people who had started but not completed primary education, and 0.22 for those who had completed their primary education, compared to people with no formal education, controlling for age group, sex, and socio-economic status (p = 0.007; d.f. 291). Crude dementia prevalence was higher in people in the lowest SES group compared to higher SES groups, but this difference was not present after controlling for age group and educational attainment. Prevalence was similar in women and men. Findings were similar in sensitivity analyses using data with an imputed values for participants with missing outcome data (**Supplementary Table 5**).

**Table 3.**
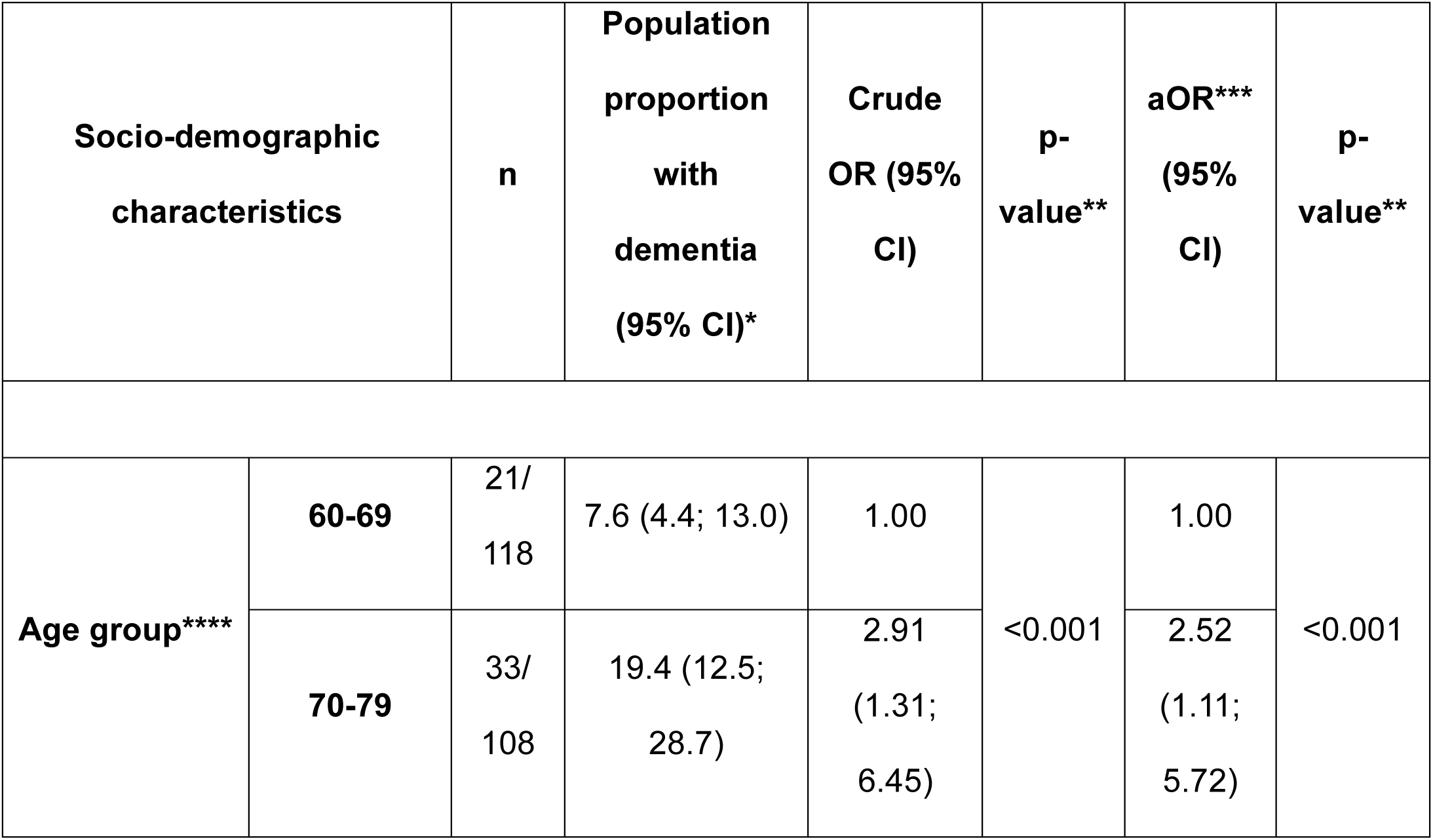
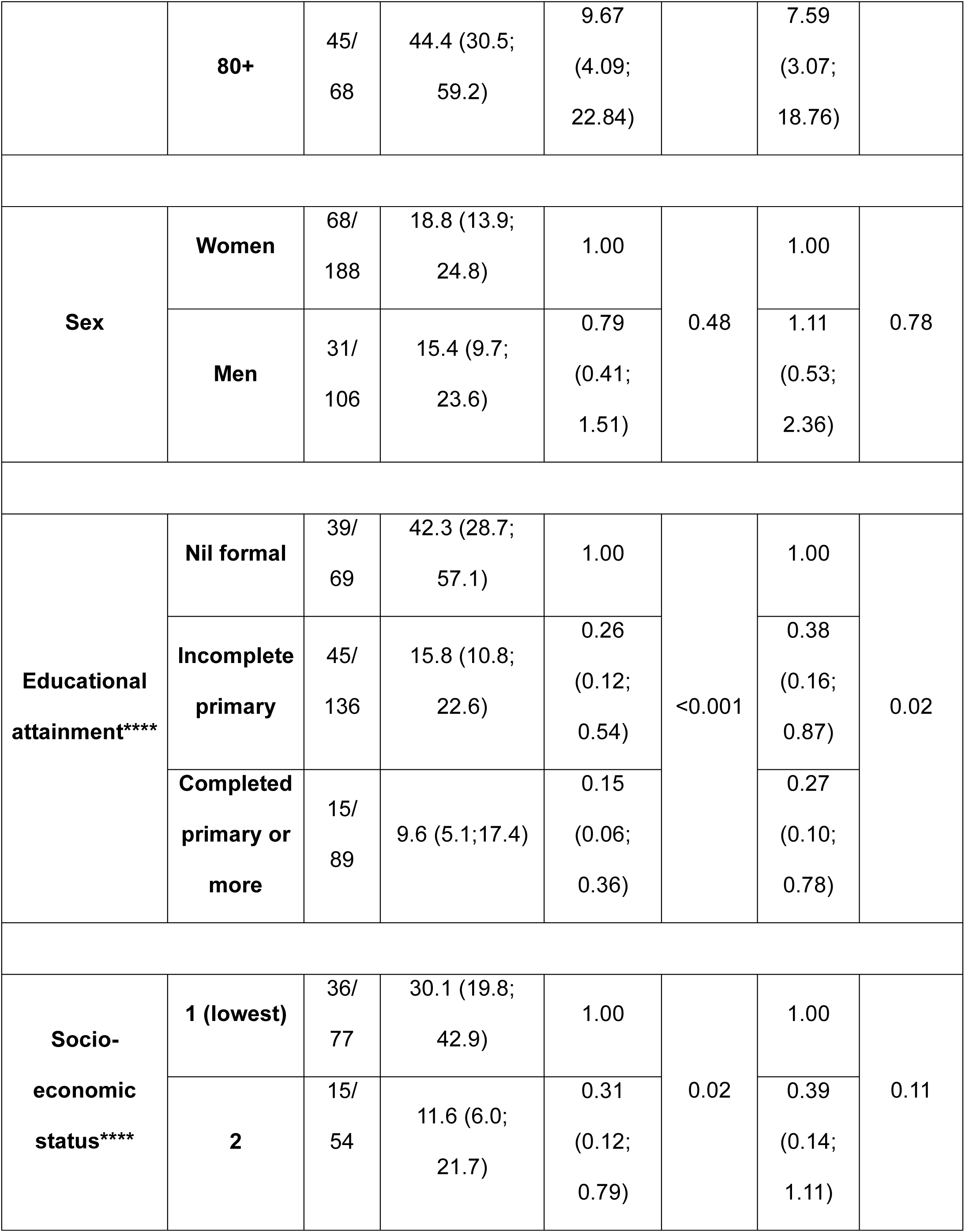
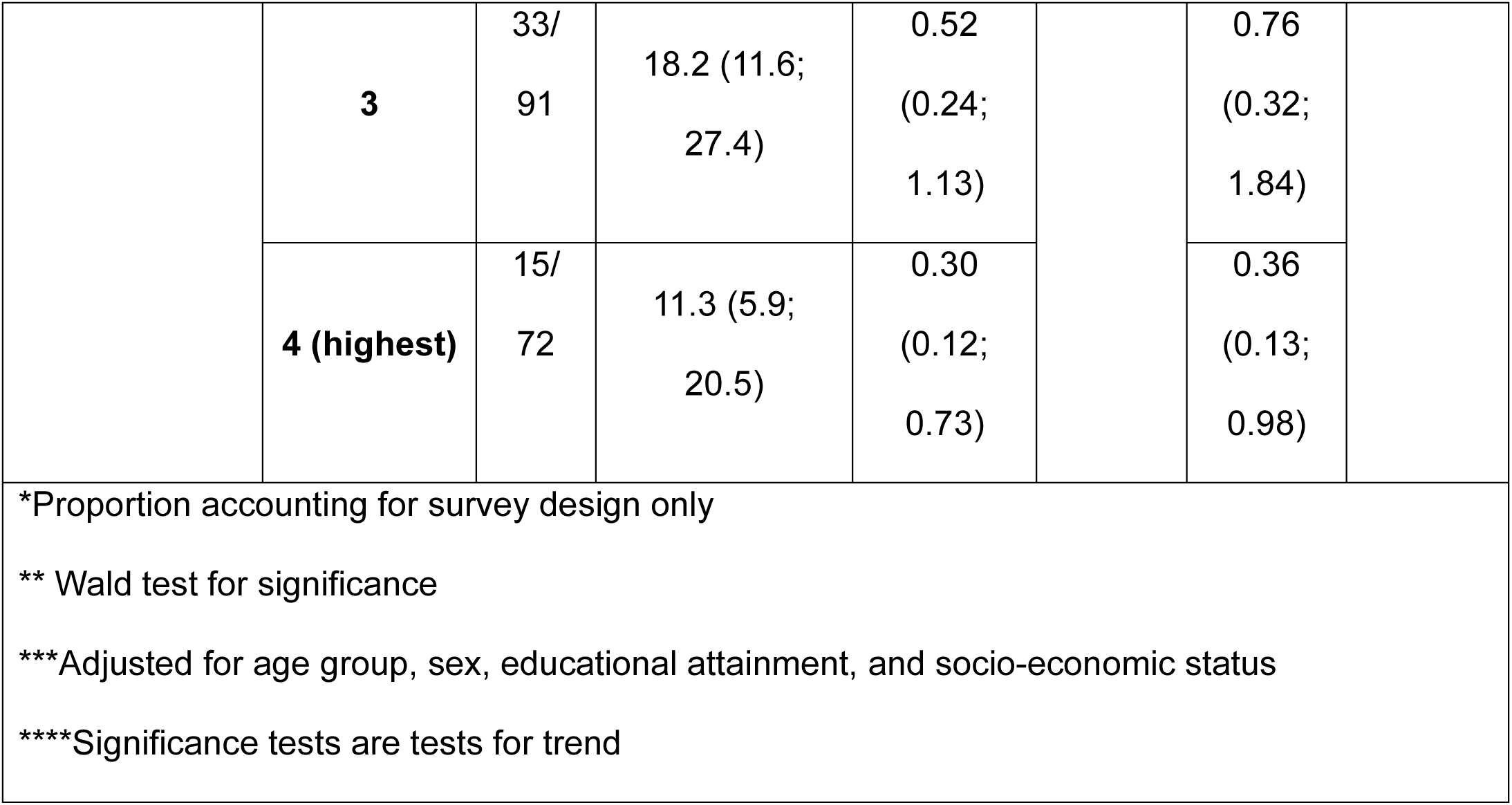
Socio-demographic correlates with probable dementia.

| Socio-demographic characteristics |  | n | Population proportion with dementia (95% CI)* | Crude OR (95% CI) | p-value** | aOR*** (95% CI) | p-value** |
| --- | --- | --- | --- | --- | --- | --- | --- |
| Age group**** | 60-69 | 21/118 | 7.6 (4.4; 13.0) | 1.00 | <0.001 | 1.00 | <0.001 |
|  | 70-79 | 33/108 | 19.4 (12.5; 28.7) | 2.91 (1.31; 6.45) |  | 2.52 (1.11; 5.72) |  |
|  | 80+ | 45/<br>68 | 44.4 (30.5;<br>59.2) | 9.67<br>(4.09;<br>22.84) |  | 7.59<br>(3.07;<br>18.76) |  |
| Sex | Women | 68/<br>188 | 18.8 (13.9;<br>24.8) | 1.00 | 0.48 | 1.00 | 0.78 |
|  | Men | 31/<br>106 | 15.4 (9.7;<br>23.6) | 0.79<br>(0.41;<br>1.51) |  | 1.11<br>(0.53;<br>2.36) |  |
| Educational attainment**** | Nil formal | 39/<br>69 | 42.3 (28.7;<br>57.1) | 1.00 | <0.001 | 1.00 | 0.02 |
|  | Incomplete primary | 45/<br>136 | 15.8 (10.8;<br>22.6) | 0.26<br>(0.12;<br>0.54) |  | 0.38<br>(0.16;<br>0.87) |  |
|  | Completed primary or more | 15/<br>89 | 9.6 (5.1;17.4) | 0.15<br>(0.06;<br>0.36) |  | 0.27<br>(0.10;<br>0.78) |  |
| Socio-economic status**** | 1 (lowest) | 36/<br>77 | 30.1 (19.8;<br>42.9) | 1.00 | 0.02 | 1.00 | 0.11 |
|  | 2 | 15/<br>54 | 11.6 (6.0;<br>21.7) | 0.31<br>(0.12;<br>0.79) |  | 0.39<br>(0.14;<br>1.11) |  |
|  | **3** | 33/<br>91 | 18.2 (11.6;<br>27.4) | 0.52<br>(0.24;<br>1.13) |  | 0.76<br>(0.32;<br>1.84) |  |
|  | **4 (highest)** | 15/<br>72 | 11.3 (5.9;<br>20.5) | 0.30<br>(0.12;<br>0.73) |  | 0.36<br>(0.13;<br>0.98) |  |

\*Proportion accounting for survey design only
\*\* Wald test for significance
\*\*\*Adjusted for age group, sex, educational attainment, and socio-economic status
\*\*\*\*Significance tests are tests for trend

**Table 4.** Comparison of disability and independence in activities of daily living and instrumental activities of daily living among people with and without dementia.

| Continuous outcome data |  |  | n | Median score** | Negative binomial regression analysis |  |  |  |
| --- | --- | --- | --- | --- | --- | --- | --- | --- |
|  |  |  |  |  | Crude count ratio | p-value | Adjusted count ratio* (95% CI) | p-value |
| WHODAS-12 Disability score | 0 - 100 (higher = more disability) | No dementia | 190 | 8.3 | 1.90 (1.45; 2.48) | <0.001 | 1.37 (0.94; 2.00) | 0.10 |
|  |  | Dementia | 92 | 30.6 |  |  |  |  |
| IDEA-IADL score | 0 – 33 (higher = less assistance needed) | No dementia | 182 | 32.0 | 0.68 (0.58; 0.80) | <0.001 | 0.77 (0.66; 0.90) | 0.001 |
|  |  | Dementia | 91 | 21.0 |  |  |  |  |

| Binary outcome data |  |  |  | n | Percentage** | Crude odds ratio (95% CI) | p-value | Adjusted odds ratio* (95% CI) | p-value |
| --- | --- | --- | --- | --- | --- | --- | --- | --- | --- |
| Care needs | Needs frequent care vs occasional care vs no care | No dementia | No care | 32/ 182 | 17.6% | 2.41 (1.07; 5.43) | 0.03 | 1.38 (0.56; 3.41) | 0.49 |
|  |  |  | Occasional care | 120/ 182 | 66.5% |  |  |  |  |
|  |  |  | Frequent care | 30/ 182 | 15.9% |  |  |  |  |
|  |  | Dementia | No care | 10/ 91 | 16.9% |  |  |  |  |
|  |  |  | Occasional care | 44/ 91 | 43.6% |  |  |  |  |
|  |  |  | Frequent care | 37/ 91 | 39.5% |  |  |  |  |
| *Adjusted for age, sex, socio-economic status and educational attainment |  |  |  |  |  |  |  |  |  |
| **Accounting for survey design |  |  |  |  |  |  |  |  |  |

Of 282 participants who attended the clinic to complete questionnaires including WHODAS-12, 5/190 participants (1.8% accounting for survey design) without dementia and 16/92 participants (11.7% accounting for survey design) with probable dementia the proxy responses were used.

We found that people with dementia had higher levels of disability, needed more assistance with instrumental activities of daily living, and had greater care needs, compared to people without dementia (**Table 5**). This was somewhat mitigated after adjustment for age, sex, socio-economic status and educational attainment, but even after adjustment, the IDEA-IADL score was, on average, 23% lower among those with dementia than those without (p=0.001, d.f.270).

**Table 5.**
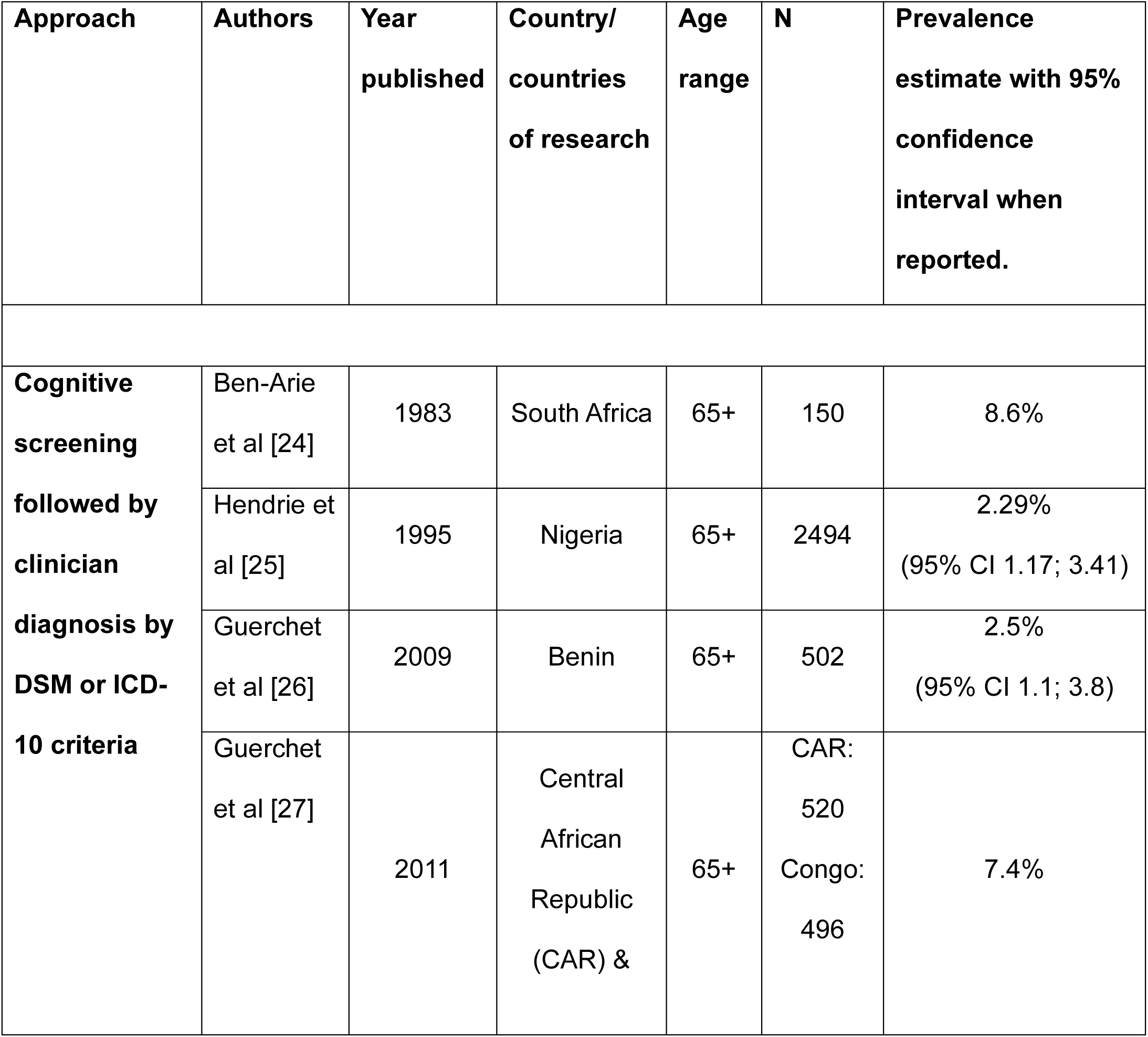
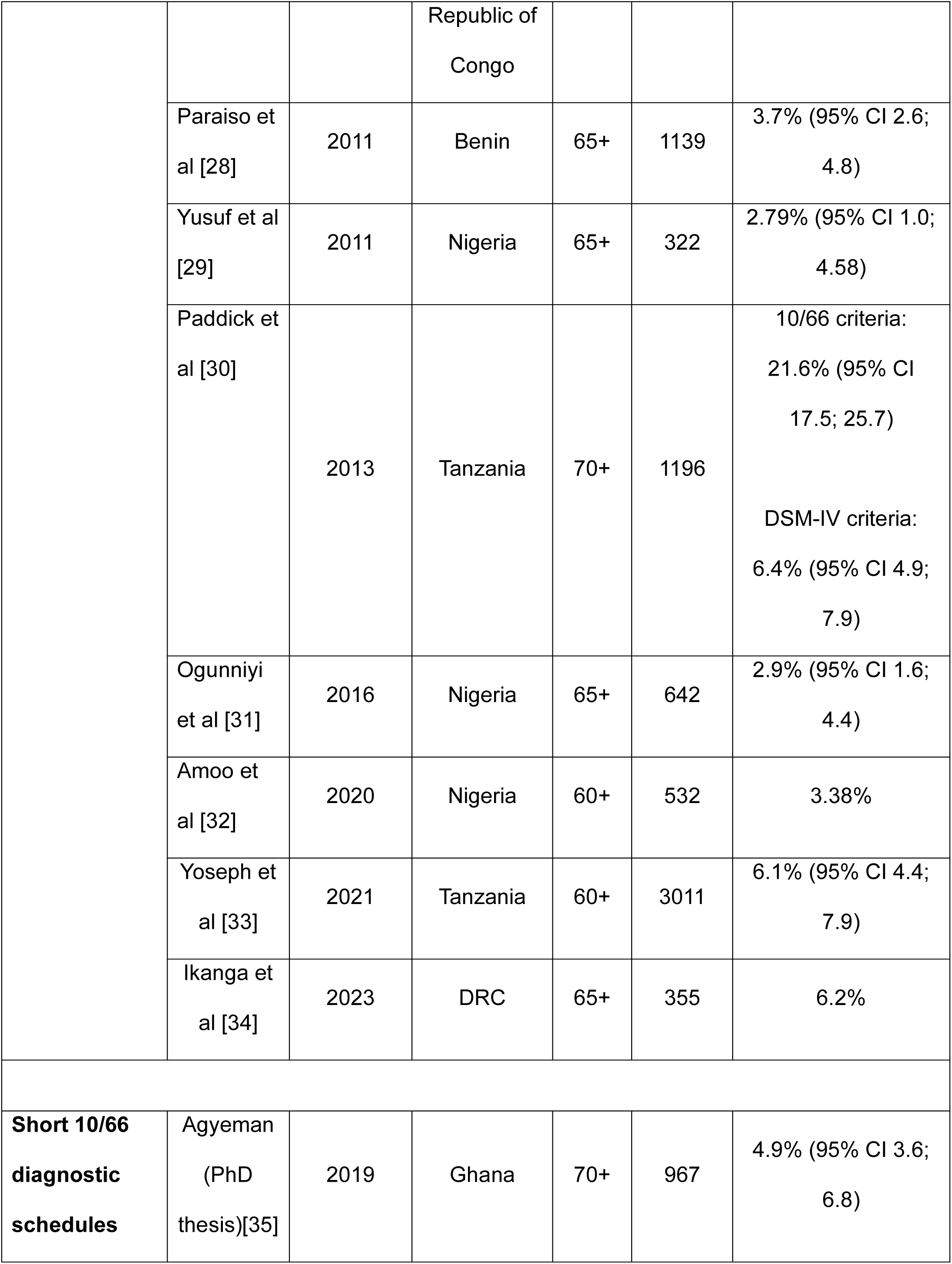
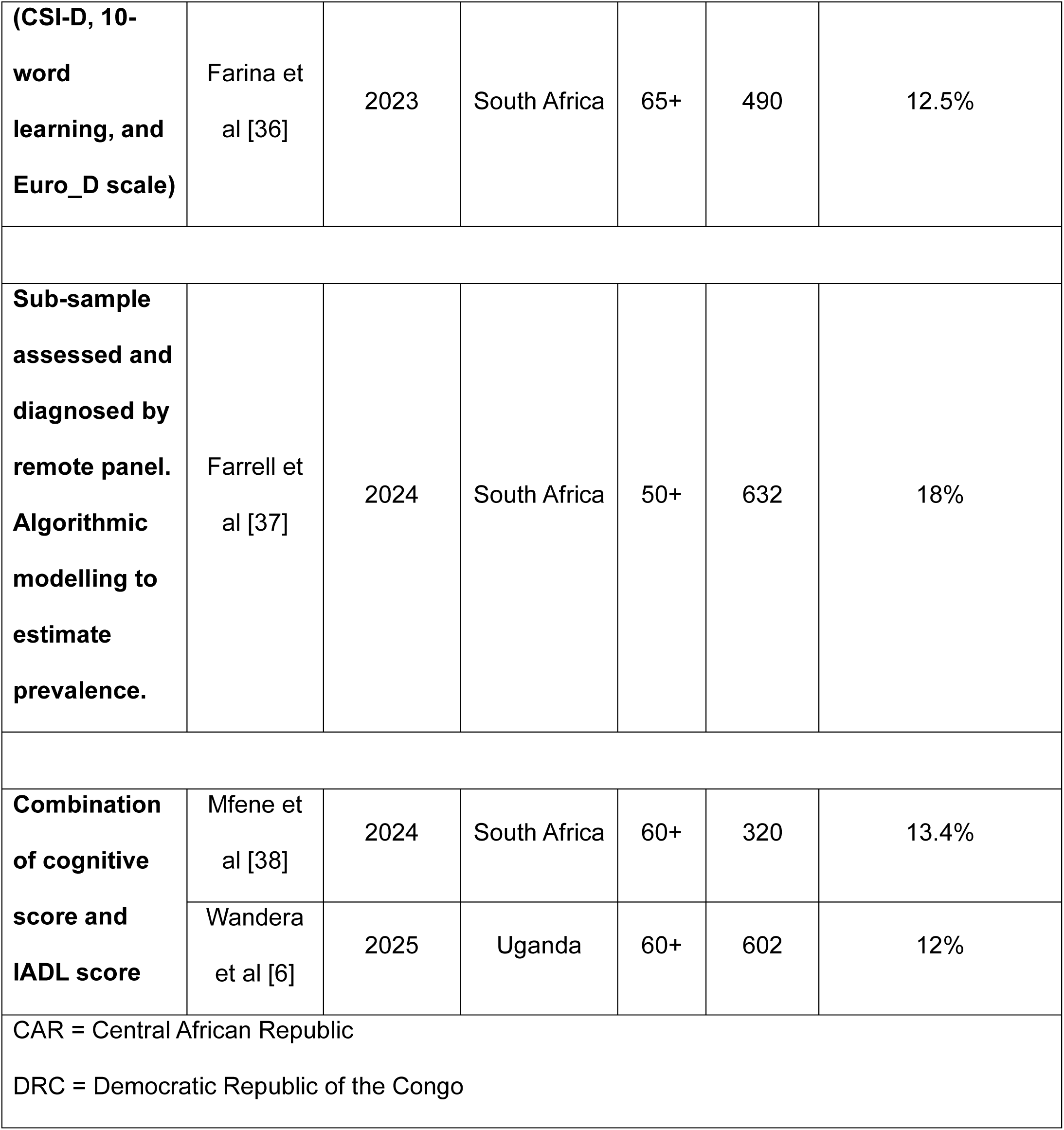
Published studies estimating dementia prevalence in African settings, categorised by methodological approach.

## Discussion

We found a high dementia prevalence of 18.6%, with most identified cases representing mild or very mild disease. Dementia was more common in the older age groups and those with lower educational attainment but was not associated with sex or socio-economic status. People experiencing dementia needed more assistance with IADLs.

The Global Burden of Disease study has estimated age-specific global dementia prevalence less than 5% in the 60-69 age group, around 10% in the 75-79 age group, rising to around 20% in the 85-89 age group.[39] Previous dementia prevalence studies in African settings have yielded a wide spread of estimates, some similar to this, and some higher, partly due to variation in study design, see **Table 5**.

Commonly, a two-phase approach is used, whereby a population of older people undergo cognitive screening, and a subgroup are assessed by a clinician using DSM criteria. The earliest of these was in Ibadan, Nigeria in 1995,[25] when dementia prevalence in adults 65 and older was estimated at 2.29% (95% CI 1.17; 3.41%). Since then, similar approaches have been used in Benin,[26] Tanzania[40], and three different sites in Nigeria,[29, 31, 32] with similar prevalence estimates obtained. This approach yielded higher estimates of 6.2 – 8.1% in urban areas in Central African Republic and Republic of Congo in 2010 and in the Democratic Republic of the Congo in 2023.[34] One criticism of this two-phase approach is that there can be substantial attrition of participants between the phases, introducing selection bias. If people with dementia are less likely to be retained, this can negatively bias the prevalence estimate.[41] A strength of our study is the high retention rate of over 90% between phases.

Other studies have used a one-phase approach, where all older people in a population undergo a single assessment that includes assessment of both cognition and function, and no clinician diagnosis is required. In recent studies in South Africa[38] and Uganda[6], teams have developed their own pragmatic algorithms based on performance at cognitive testing and informant responses to IADL questionnaires, and have estimated dementia prevalence in people aged 60+ as 13.4%[38] and 12.0%[6] respectively.

In a subset of the HAALSI cohort in South Africa, dementia diagnosis was made remotely by an expert panel following assessment with the Harmonised Cognitive Assessment Protocol (HCAP). Algorithmic modelling was then used to estimate a dementia prevalence within the whole cohort of 18% (95% CI 15; 22%).[37]

Two studies have used a shortened version of the 10/66 diagnostic criteria, where the much shorter Euro-D is substituted for the GMS, and prevalence was estimated as 4.9% in adults aged 70+ in Ghana and 2.5% in adults aged 65+ in South Africa.[36]

It has been argued that the DSM criteria may underestimate dementia prevalence, particularly in lower income settings, partly due to its reliance on informant report of functional impairment.[23] In some lower income settings, cognitive impairment may not have the same impact on function in the early stages, due to higher levels of baseline support from family and community members, and there may be differences in reporting behaviour of informants around functional impairment.[23]

In the Hai District of Tanzania, prevalence according to DSM-IV criteria was estimated at 6.4% (4.8; 7.9%) with a higher age cut-off of 75 years.[40] However, when they applied the 10/66 diagnostic criteria to the same population and estimated a much higher prevalence of 21.6% (95% CI 17.5; 25.7%), similar our findings of 19.4% in people aged 70-79 and 44.4% in those aged 80+. This finding was in keeping with those of the multi-site 10/66 Dementia Research Group, who demonstrated that while their prevalence estimates for 10/66 dementia were consistently substantially higher than their estimates for DSM-IV dementia. However, the authors of the 10/66 study also noted that participants who were 10/66 dementia positive but DSM-IV dementia negative were indistinguishable from DSM-IV dementia patients in terms of disability, supporting the notion that the 10/66 approach was identifying additional true cases over the DSM-IV approach.[23]

Our prevalence estimate of 18.6% in adults aged 60 and over is comparable to the 10/66 results found in Hai, Tanzania,[30] and to the algorithmically derived results from South Africa[37] but higher than most other estimates in the continent. Interestingly, we found that 87.6% of people with probable dementia were categorised as mild or very mild using the CDR. One interpretation is that the 10/66 algorithm is insufficiently specific in this population, and some of the participants with CDR 0.5 (very mild or questionable dementia) may have been misclassified, particularly if they had received lower levels of formal education. However, people with probable dementia showed persistently lower levels of independence in IADLs than those without dementia, which does suggest that impaired cognition is affecting their higher-level function, even if their symptoms have not yet progressed to experiencing disability or needing support with ADLs (demonstrated by a need for care). It is also important to note that our construction of a CDR score was through extraction of individual data points that had been collected as part of the 10/66 dementia diagnostic approach, and not as part of a semi-structured clinical interview as CDR was designed to be used,[42] which may affect its validity.

We did not identify any participant with severe dementia. It may be that there are people with severe dementia in the community, but they were missed in our data collection processes. This is more likely to have happened at Phase 1 than Phase 2, where we saw higher non-participation rates in the oldest old participants, compared to the recruitment for Phase 2, where recruitment was similar across age and cognitive function groups. Additionally, mortality rates are consistently higher for people with dementia than those without, particularly in lower income settings,[43] and among those with more severe dementia.[44]. Paddick et al found that people identified with dementia of any severity in Tanzania were more than 6 times more likely to die within 4 years of follow-up than people with unimpaired cognition.[45] This limits the identification of people with severe dementia in cross-sectional studies.

We found that dementia was associated with lower educational attainment. While the protective effect of higher levels of education is well-documented, this is often based on analyses using incomplete secondary education as a baseline of low education.[46] We found a significantly reduced odds of dementia with even incomplete primary education compared to no formal education. This has also been seen in some other studies in African settings,[47, 48] suggesting that even very low levels of education may confer some cognitive protection, either directly or through downstream effects on lifestyle exposures.

A key strength of this study is the very high recruitment and retention rates, limiting the risk of selection bias. This was achieved through working within an established population cohort with a strong relationship with the community and embedded community outreach strategies. We were also able to facilitate participation by visiting participants at home for their cognitive and mental health assessment, and by providing transport to the clinic for the additional data collection procedures. Furthermore, as the study was embedded in a fully enumerated population, we had background information on all census participants, including those who did not participate in either phase of the study. This allowed us to estimate dementia prevalence within the whole underlying population, whilst being aware of the assumptions that this required. We were therefore able to calculate a prevalence estimate using both a complete case analysis and as applied to the whole population. However, it is possible that both these estimates are biased if people with more severe dementia were likely to be missed, which could happen if their family was sheltering them or they themselves were declining to participate. This would have led to an under-estimate of dementia prevalence. Conversely, people without dementia may have been more likely to be missed if they were engaged in commercial or caretaking activities and had less time to spare, leading to an over-estimate of dementia prevalence.

We chose to use the 10/66 algorithm to classify cases of dementia, as it is unique in its rigorous validation against clinician diagnosis in multiple low- and middle-income settings. However, it is worth considering that most of the validation sites were in Latin America and Asia, with little data from Africa.[8] Performance in cognitive tests, mental health presentations, and informant impressions of a participant’s function are influenced by culture and context, and it may be that the 10/66 approach has a lower accuracy here than other settings. Nonetheless, the approach has advantages, particularly a standardised data collection process that does not require a clinician diagnosis, increasing feasibility to roll out at scale in lower resource settings, and the direct comparability of results with other 10/66 sites. Deeper phenotyping of participants including with biomarkers of Alzheimer’s and vascular pathology would help validate the clinical findings and indicate underlying pathological processes leading to dementia in this population.

## Conclusion

We found that dementia prevalence in older adults in rural Uganda was high – almost one in five adults aged 60 and over. Even though most cases were mild, people with dementia needed significantly more support with instrumental activities of daily living than people without dementia.

Any health and social care services planned for older people in rural Uganda must be dementia-friendly and accessible and include support for people with dementia and their families.

## Data Availability

All data produced in the present study are available upon reasonable request to the authors.

## Abbreviations

ADL: Activities of daily living
CDR: Clinical dementia rating
CSI-D: Community screening instrument for dementia
d.f.: Degrees of freedom
DSM: Diagnostic and statistical manual of mental disorders
GMS: Geriatric Mental State
GPC: General Population Cohort
HCAP: Harmonised Cognitive Assessment Protocol
IADL: Instrumental activities of daily living
ICD: International classification of diseases
IDEA: Intervention for Dementia in Elderly Africans
MAR: Missing at random
MICE: Multiple imputation chained equations
MNAR: Missing not at random
OR: Odds ratio
SAGE: Study on global AGEing and adult health
SD: Standard deviation
SES: Socioeconomic status
UBACC: University of California, San Diego Brief Assessment of Capacity to Consent
WHODAS: WHO Disability Assessment Schedule
WOPS: Wellbeing of Older People’s Study

## Acknowledgements

We acknowledge and thank the participants and their caregivers throughout this study.

