## Supplementary material for "Prevalence of dementia among community-dwelling older adults in rural Uganda – findings from DEPEND Uganda (Dementia Epidemiology, unmet need and co-developing solutions in Uganda)"

**Supplementary Materials**

*Supplementary table 1. Socio-demographic predictors of non-participation in the Medical Survey, among the census population.*

| Exposures | | n/N (%) non-participation in the Medical Survey | Crude OR of non-participation | p-value* | Adjusted OR of non-participation | p-value* |
|---|---|---|---|---|---|---|
| **Sex** | Female | 153 / 984 (15.6) | 1 | 0.28 | 1 | 0.15 |
| | Male | 111 / 631 (17.6) | 1.16 (0.89; 1.52) | | 1.23 (0.93; 1.63) | |
| **Age group** | 60-69 | 139 / 968 (14.4) | 1 | <0.001 | 1 | <0.001 |
| | 70-79 | 68 / 417 (16.3) | 1.16 (0.85; 1.59) | | 1.17 (0.85; 1.61) | |
| | 80-89 | 42 / 192 (21.9) | 1.67 (1.1; 2.46) | | 1.71 (1.14; 2.54) | |
| | 90+ | 15 / 38 (39.5) | 3.89 (1.98; 7.64) | | 3.67 (1.83; 7.38) | |
| **Formal education** | Nil | 49 / 250 (19.6) | 1.26 (0.87; 1.82) | 0.25 | 1.17 (0.80; 1.73) | 0.26 |
| | Less than primary | 120 / 739 (16.2) | 1 | | 1 | |
| | Completed primary | 46 / 310 (14.8) | 0.90 (0.62; 1.30) | | 0.94 (0.64; 1.38) | |
| | Less than completed secondary | 26 / 173 (15.0) | 0.91 (0.58; 1.45) | | 0.86 (0.54; 1.37) | |
| | Completed secondary | 19 / 91 (20.9) | 1.36 (0.79; 2.34) | | 1.37 (0.78; 2.42) | |
| | Tertiary | 4 / 52 (7.7) | 0.43 (0.15; 1.21) | | 0.41 (0.14; 1.18) | |
| **SES** | 1 | 56 / 379 (14.8) | 1 | 0.72 | 1 | 0.58 |
| | 2 | 41 / 240 (17.1) | 1.19 (0.77; 1.84) | | 1.24 (0.79; 1.93) | |
| | 3 | 94 / 538 (17.5) | 1.22 (0.85; 1.75) | | 1.29 (0.89; 1.87) | |
| | 4 | 73 / 458 (15.9) | 1.09 (0.75; 1.60) | | 1.22 (0.82; 1.82) | |
| *Wald test for significance | | | | | | |

*Supplementary Table 2. Socio-demographic predictors of non-participation in DEPEND, among the sampled participants*

| Exposures | | n/N non response in the DEPEND study (%) | Crude OR of non-response | p-value* | Adjusted OR of non-response** | p-value* |
|---|---|---|---|---|---|---|
| **Sex** | Female | 22/210 (10.5) | 1 | 0.20 | 1 | 0.23 |
| | Male | 7/113 (6.2) | 0.56 (0.23; 1.36) | | 0.58 (0.24; 1.41) | |
| **Age group** | 60-69 | 17/150 (11.3) | 1 | 0.05 | 1 | 0.06 |
| | 70-79 | 2/105 (1.9) | 0.15 (0.03; 0.67) | | 0.15 (0.04; 0.67) | |
| | 80-89 | 8/53 (15.1) | 1.39 (0.56; 3.44) | | 1.35 (0.54; 3.34) | |
| | 90+ | 2/15 (13.3) | 1.20 (0.25; 5.80) | | 1.15 (0.24; 5.55) | |
| **Formal education** | Nil | 9/78 (11.5) | 1.36 (0.56; 3.35) | 0.92 | 1.48 (0.55; 4.00) | 0.94 |
| | Less than primary | 13/149 (8.7) | 1 | | 1 | |
| | Completed primary | 4/46 (8.7) | 1.00 (0.31; 3.22) | | 1.09 (0.32; 3.69) | |
| | Less than completed secondary | 2/33 (6.1) | 0.67 (0.14; 3.15) | | 0.80 (0.17; 3.86) | |
| | Completed secondary | 1/12 (8.3) | 0.95 (0.11; 7.96) | | 1.20 (0.13; 10.75) | |
| | Tertiary | 0/5 (0) | n/a | | n/a | |
| **SES** | 1 | 10/87 (11.5) | 1 | 0.34 | 1 | 0.43 |
| | 2 | 1/55 (1.8) | 0.14 (0.02; 1.15) | | 0.17 (0.02; 1.42) | |
| | 3 | 10/101 (9.9) | 0.85 (0.33; 2.14) | | 0.93 (0.36; 2.40) | |
| | 4 | 8/80 (10.0) | 0.86 (0.32; 2.28) | | 0.96 (0.35; 2.62) | |
| **Cognitive screen** | Unimpaired | 14/169 (8.3) | 1 | 0.12 | 1 | 0.15 |
| | Mildly impaired | 4/78 (5.1) | 0.60 (0.19; 1.88) | | 0.65 (0.19; 2.16) | |
| | Impaired | 11/77 (14.3) | 1.87 (0.81; 4.34) | | 2.01 (0.77; 5.25) | |

*Wald test
**Adjusted for age and sex

*Additional information about construction of socio-economic status variable*

Socio-economic indicators included in the census were characteristics of the participant's dwelling (e.g. building materials of the roof and state of repair), ownership of durable assets (e.g. tables, bicycle, television), ownership of livestock, access to certain services (e.g. electricity), and home ownership. We created a single variable as a proxy for socio-economic status, using a Mokken analysis to determine which variables to include. Variables used to make up the final variable were floor material (mud versus cement or tiles), ownership of a mobile phone, chair, table, television, clock, car and refrigerator, and whether any household members owned a property to rent. We found that this to be a unidimensional, monotonic and hierarchical scale.

*Supplementary table 3. Variables included in socio-economic status variable, mean score and Loevinger H coefficient at Mokken analysis*

| **Variable*** | **Mean score** | **Loevinger H coefficient** |
|---|---|---|
| Mobile phone ownership | 0.8705 | 0.65252 |
| Chair ownership | 0.6183 | 0.59017 |
| Table ownership | 0.5607 | 0.61445 |
| TV ownership | 0.3476 | 0.64449 |
| Motorbike ownership | 0.1983 | 0.41867 |
| Ownership of another property which is rented out | 0.0706 | 0.43133 |
| Clock ownership | 0.0657 | 0.53364 |
| Car ownership | 0.0229 | 0.36812 |
| Refrigerator ownership | 0.0173 | 0.54937 |
| Overall | - | 0.55358 |

*Supplementary table 4. Prevalence of probable dementia, using complete case analysis and not accounting for differential non-response by age group*

| | **n** | **% (95% CI)*** |
|---|---|---|
| Screen negative | 15/155 | 9.7% (5.9%; 15.5%) |
| Screen positive for mildly impaired cognition | 33/74 | 44.6% (33.5; 56.3%) |
| Screen positive for impaired cognition | 51/65 | 78.5% (66.5%; 87.0%) |
| **Population prevalence back-weighted for sampling probability only.** | **99/294** | **17.4% (13.6; 22.0%)** |

*Supplementary Table 5. Socio-demographic correlates with 10/66 dementia, accounting for missing data through multiple imputation of the dementia outcome variable, and inverse-probability weighting to adjust for missingness by age group at the medical survey level.*

| Socio-demographic characteristics | | n | Percentage with dementia (95% CI)* | aOR*** (95% CI) | p-value |
|---|---|---|---|---|---|
| **Age group**** ** | **60-69** | 21/ 118 | 7.6 (3.3 ;11.9) | 1.00 | <0.001 |
| | **70-79** | 33/ 108 | 19.4 (11.3; 27.5) | 2.53 (1.12; 5.74) | |
| | **80+** | 45/ 68 | 44.8 (29.7; 60.0) | 7.71 (3.12; 19.03) | |
| **Sex** | **Women** | 68/ 188 | 20.1 (14.3; 25.9) | 1.00 | 0.78 |
| | **Men** | 31/ 106 | 16.2 (9.3; 23.1) | 1.11 (0.52; 2.35) | |
| **Educational attainment******** | **Nil formal** | 39/ 69 | 42.9 (28.8; 57.1) | 1.00 | 0.03 |
| | **Incomplete primary** | 45/ 136 | 17.1 (10.7; 23.4) | 0.38 (0.17; 0.89) | |
| | **Completed primary or more** | 15/ 89 | 10.1 (4.0; 16.2) | 0.28 (0.10; 0.81) | |
| **Socio-economic status******** | **1 (lowest)** | 36/ 77 | 31.2 (19.8; 42.6) | 1.00 | 0.18 |
| | **2** | 15/ 54 | 12.1 (4.5; 19.6) | 0.40 (0.14; 1.12) | |
| | **3** | 33/ 91 | 19.7 (11.3; 28.1) | 0.77 (0.32; 1.85) | |
| | **4 (highest)** | 15/ 72 | 12.4 (4.9; 19.8) | 0.36 (0.13; 0.98) | |

*Proportion accounting for survey design and missingness
**Wald test
***Adjusted for age group, sex, educational attainment, and socio-economic status
****Significance tests are tests for trend

*Supplementary table 6. Socio-demographic correlates with 10/66 dementia of clinical dementia rating 1 or more*

| Socio-demographic characteristics | | n | Percentage with dementia and CDR 1+ (95% CI)* | aOR (95% CI)*** | p-value** |
|---|---|---|---|---|---|
| **Age group******** | **60-69** | 10/ 118 | 2.9% (1.2; 6.8%) | 1.00 | <0.0001 |
| | **70-79** | 16/ 108 | 8.1% (4.2 ;15.1%) | 2.86 (0.94; 7.75) | |
| | **80+** | 31/ 68 | 27.0% (16.9; 40.3%) | 10.91 (3.44; 34.55) | |
| **Sex** | **Women** | 38/ 188 | 9.6% (6.3; 14.4%) | 1.00 | 0.80 |
| | **Men** | 19/ 106 | 6.7% (3.6; 12.2%) | 0.89 (0.36; 2.19) | |
| **Educational attainment******** | **Nil formal** | 19/ 69 | 15.9% (8.9%; 27.0%) | 1.00 | 0.25 |
| | **Incomplete primary** | 31/136 | 10.0% (6.2; 15.6%) | 0.97 (0.39; 2.40) | |
| | **Completed primary or more** | 7/ 89 | 3.9% (1.4; 10.1%) | 0.47 (0.13; 1.77) | |
| **Socio-economic status******** | **1 (lowest)** | 20/ 77 | 12.3% (7.2; 20.4%) | 1.00 | 0.57 |
| | **2** | 8/ 54 | 6.7% (2.5; 17.1%) | 0.67 (0.17; 2.62) | |
| | **3** | 20/ 91 | 9.3% (5.2%; 16.0%) | 1.05 (0.36; 3.04) | |
| | **4 (highest)** | 9/ 72 | 5.9% (2.4%; 13.7%) | 0.58 (0.18; 1.92) | |

*Proportion accounting for survey design only
**Wald test
***Adjusted for age group, sex, educational attainment, and socio-economic status
****Significance tests are tests for trend
